# Seasonality in living kidney donation in the United States from 1995-2019

**DOI:** 10.1101/2022.03.11.22271849

**Authors:** Andrew Arking, Gabriella Kaddu, Dorry L. Segev, Jacqueline Garonzik-Wang, Abimereki D. Muzaale, Fawaz Al Ammary

**Affiliations:** Department of Surgery, Johns Hopkins University School of Medicine, Baltimore, MD; Department of Epidemiology, Johns Hopkins School of Public Health, Baltimore, MD; Scientific Registry of Transplant Recipients, Minneapolis, MN; Department of Medicine, Johns Hopkins University School of Medicine, Baltimore, MD

**Keywords:** Living Kidney Donors, Seasonality, Barriers

## Abstract

For nearly two decades, the annual number of US living kidney donors has been characterized by worrying patterns of decline and no factors have been identified to explain and reverse these patterns. Evidence suggests that there is seasonality in living kidney donation; herein we investigate whether potentially modifiable social, economic, and structural issues might explain this seasonality. Using donor-registry data from the Scientific Registry of Transplant Recipients, we described this seasonality in living kidney donation and used Poisson regression stratified by both donor–recipient biological relationship and estimated household income tertile to quantify these trends. In every decade from 1980-2020, there was a summer-only surge in living kidney donations (13%-25% for biologically related donors and 10%-17% for unrelated donors). This summer-only surge was evident for the months of June, July, and August when compared with January for each given year and statistically significant in some groups (range of incidence rate ratio [IRR] for related donors: 1.05-1.34; IRR for unrelated donors: 1.08-1.19). We observed this summer-only surge across all three income tertiles ($73,544+, $52,635- $73,544, and <$52,635) and regardless of donor-recipient relationship. Seasonal variation in donation is associated with structural factors, which may serve as potential targets for interventions to increase donation.

## INTRODUCTION

The narrative in all previously described changes in number of live kidney donors in the US is a narrative of annual trends in live kidney donation. Within this approach, there was exponential growth in donation from 1987-2004 in part attributable to the adoption of laparoscopic donor nephrectomy after 1995 [1]. Following this period of growth, there was a leveling-off and an actual decline in donation from 2005-2017 [2]. In these past 15 years, no attributable cause of annual decline has been proposed. What is known is that the decline was most clearly observed among donors who were biologically related to the recipient. And between 2017-2019, the first “sustained” increase in live kidney donation in 15 years has been reported, driven chiefly by an increase in unrelated donors [3].

These annual trends exist in parallel to seasonal trends wherein patterns of kidney donation fluctuate consistently within each year, independent of annual trends. An increase of donations during summer months—June, July, and August—has been previously identified but without any further investigation or quantification [4]. Understanding the role of seasonal trends within the landscape of kidney donation in the United States could alter the framework through which we investigate overall trends in donation by adding a novel effect modifier to analyses. Notably, establishing the contribution of seasonal trends could provide valuable insight into structural barriers which potential donors face and point to potential policy changes to enable more potential donors to donate.

In this study, we assessed whether the changes in the number of donors in the US are better understood by studying seasonal fluctuations in donation in fall, winter, spring, and summer months than by assessing annual trends, which no longer permit a clear and actionable narrative. We used linear mixed models to formally test whether seasonal variation in donation better explains recent changes in donation than hitherto described annual trends in donation. We subsequently assessed how these seasonal variations have changed over time and investigated if donors from lower income households display different seasonal trends from donors from higher income households.

## METHODS

### Study Population

This study used data from the Scientific Registry of Transplant Recipients (SRTR). The SRTR data system includes data on all donor, wait-listed candidates, and transplant recipients in the US, submitted by the members of the Organ Procurement and Transplantation Network (OPTN). The Health Resources and Services Administration (HRSA), U.S. Department of Health and Human Services provides oversight to the activities of the OPTN and SRTR contractors. This dataset has previously been described elsewhere [5]. Through this reporting, 147,965 adult live donors between October 1, 1987, and August 30, 2019, were included in this study. This Institutional Review Board of Johns Hopkins University has determined that this study qualifies as exempt research under the DHHS regulations because it uses deidentified data.

### Trends in Living Kidney Donation

We stratified the donor population into related and unrelated groups because each group has different demographics and historical trends [2, 3]. Donor-recipient relationship was ascertained by self-report at donor evaluation. Paired donors were excluded because their small sample size (N=6975) did not provide statistical power to make inferences. We further divided donors into income tertiles by associating donor home zip code with median household income as reported by the US Census Bureau in the 5-year estimate in the 2018 American Community Survey [6]. For certain analyses, to better understand historical trends in donation, we grouped the donors by decade of donation. The outcome of interest was the change in live kidney donation per month within and between years.

To assess the contributions of within-year and between-year variation to changes in living kidney donation, we performed a linear mixed-effects model for number of donors per month and year. Intraclass correlation (ICC), which reflects the consistency of the monthly data within each year, was determined based on the model’s coefficients for between-year variation (τ) and within-year variation (σ) using the formula τ^2^ / (τ^2^ + σ^2^). Due to previously shown differences between trends in donation prior to 2005 and after 2005, we also determined these values separately for these two time periods.

### Within-Year Seasonal Variation in Living Kidney Donors

We used Poisson regression to estimate the change in number of donors between months (incidence rate ratio, IRR). The IRR indicates the proportional decline or increase in the number of donors between months, using January as the reference. IRRs were also estimated by dividing the monthly data into seasons—Spring, Summer, Fall, Winter—to help discern trends throughout the year. To adjust for the different lengths of months, we normalized case number based on the number of days in the month, using the formula: (Number of cases) / (Number of days in that month) × (Average number of days in a month, 365.25/12) [4].

All analyses were performed using RStudio Version 1.3.959 for Windows (RStudio, PBC) running R 4.0.2. All hypothesis tests were 2 sided (α = 0.05)

## RESULTS

### Study Population

Among 147,965 live kidney donors, characteristics remained consistent across the season of donation but varied between related and unrelated donors. For related donors, who comprised two-thirds of total donors, median age at donation was 39 years; 57% were female, 69% were white, 15% were Black, and 15% were Hispanic. Eighty-three percent of donors were younger than 50 and 17% were older than 50 years old. No donors had diabetes at baseline, but 1% had hypertension. Median systolic/diastolic blood pressure was 120/74 mm Hg, median body mass index was 27 kg/m^2^, 25% had a history of smoking cigarettes, 15% graduated from college, and 6% had postgraduate education. Thirty-one percent of donors were in the highest income tertile ($73,321 and over), 33% were in the middle tertile ($52,463- $73,321), and 36% were in the lowest tertile ($52,463 and lower) Their median year of donation was 2004.

Unrelated donors, who comprised one-third of total donors, tended to be older, more frequently white, wealthier, and more educated. Median age at donation was 44 years; 65% were female, 82% were white, 8% were Black, and 10% were Hispanic. Seventy-one percent of donors were younger than 50 and 29% were older than 50 years old. None of the donors had diabetes at baseline, but 2% had hypertension. Median systolic/diastolic blood pressure was 120/74 mm Hg, median body mass index was 27 kg/m^2^, 25% had a history of smoking cigarettes, 22% graduated from college, and 11% had postgraduate education. Thirty-seven percent of donors were in the highest income tertile (over $73,544), 33% were in the middle tertile ($52,635-$73,544), and 29% were in the lowest tertile (under $52,635). Unrelated donors are a more recent phenomenon, with a median year of donation of 2009 (Table 1).

**Table 1.** Baseline characteristics of live kidney donors in the United States in summer (June, July, August) and non-summer months in 1987-2020, stratified by donor type (related, unrelated) Abbreviations: IQR: interquartile range; mmHg: millimeters of mercury; SBP, systolic blood pressure; DBP: diastolic blood pressure; BMI: body mass index; HS: high school. 44% related education missing, 22% unrelated Note others for race Creat, bmi, htn Hypertension was defined as predonation documented use of antihypertensive therapy/history of hypertension (% missing)

|  | Related |  | Unrelated |  |
| --- | --- | --- | --- | --- |
|  | Summer<br>N=29674 | Non-Summer<br>N=76351 | Summer<br>N=14308 | Non-Summer<br>N=39297 |
| Age, Median [IQR] | 38 [30-47] | 38 [30-47] | 44 [35-52] | 44 [35-52] |
| Age Range, % |  |  |  |  |
| 18-39 | 57.4 | 57.1 | 39.6 | 39.9 |
| 40-49 | 26.7 | 26.1 | 31.4 | 30.9 |
| 50+ | 15.9 | 16.8 | 29 | 29.2 |
| Recipient Age, Median [IQR] | 41 [27-54] | 42 [29-55] | 49 [39-58] | 49 [39-58] |
| Year of Donation, Median [IQR] | 2004 [1998-2010] | 2004 [1998-2011] | 2009 [2004-2015] | 2010 [2004-2015] |
| Female, % | 59 | 56.3 | 64.9 | 63.6 |
| Blood Pressure, mmHg [IQR] |  |  |  |  |
| SBP | 120 [111-130] | 120 [112-130] | 120 [111-130] | 120 [112-130] |
| DBP | 74 [68-80] | 74 [68-80] | 74 [68-80] | 74 [68-80] |
| BMI, Median [IQR] | 27 [24-30] | 27 [24-30] | 27 [24-29] | 26 [24-30] |
| Hypertension, % | 1.2 | 1.3 | 2.6 | 2.7 |
| Diabetes, % | 0 | 0 | 0 | 0 |
| Smoke, % | 23.9 | 25.4 | 24.1 | 24.9 |
| Race, % |  |  |  |  |
| Black | 14.8 | 14.6 | 7.9 | 7.6 |
| Hispanic | 15.5 | 15.5 | 9.9 | 9.7 |
| White | 69.8 | 69.9 | 82.2 | 82.7 |
| Income Tertile, % |  |  |  |  |
| 1st (\$73,544+) | 32.1 | 31.5 | 36.4 | 37.4 |
| 2nd (\$52,635-\$73,544) | 32.5 | 32.9 | 33.8 | 33.3 |
| 3rd (\$0-\$52,635) | 35.4 | 35.6 | 29.8 | 29.4 |
| Highest Education Level, % |  |  |  |  |
| <= HS | 18.1 | 19.3 | 21 | 21.3 |
| Attended College | 15.3 | 15.6 | 20.9 | 21.1 |
| College Graduate | 15.6 | 15.5 | 23.1 | 23.6 |
| Post College | 6.2 | 5.9 | 12.7 | 11.5 |
| Not reported | 44.8 | 43.6 | 22.4 | 22.5 |

### Trends in Living Kidney Donation

Kidney donations increased monotonically between 1987 and 2004 and declined thereafter before experiencing a resurgence starting in 2017. Notably, within-year seasonal variance increased after 2004 (Figure 1). Prior to 2005, between-year variance was 169 donations/month and within-year variance was 42 donations/month, yielding an intraclass correlation of 0.94. In contrast from 2005 onward, between-year variance was 32 donations/month and within-year variance was 52 donations/month, yielding an intraclass correlation of 0.28. These different intraclass correlations indicate that between-year variation explains trends in donation prior to 2005 well, but does not do so for trends since 2005. Intra-year variation (sigma) increased by 30%, from 40 to 52, between these time periods.

**Figure 1.**
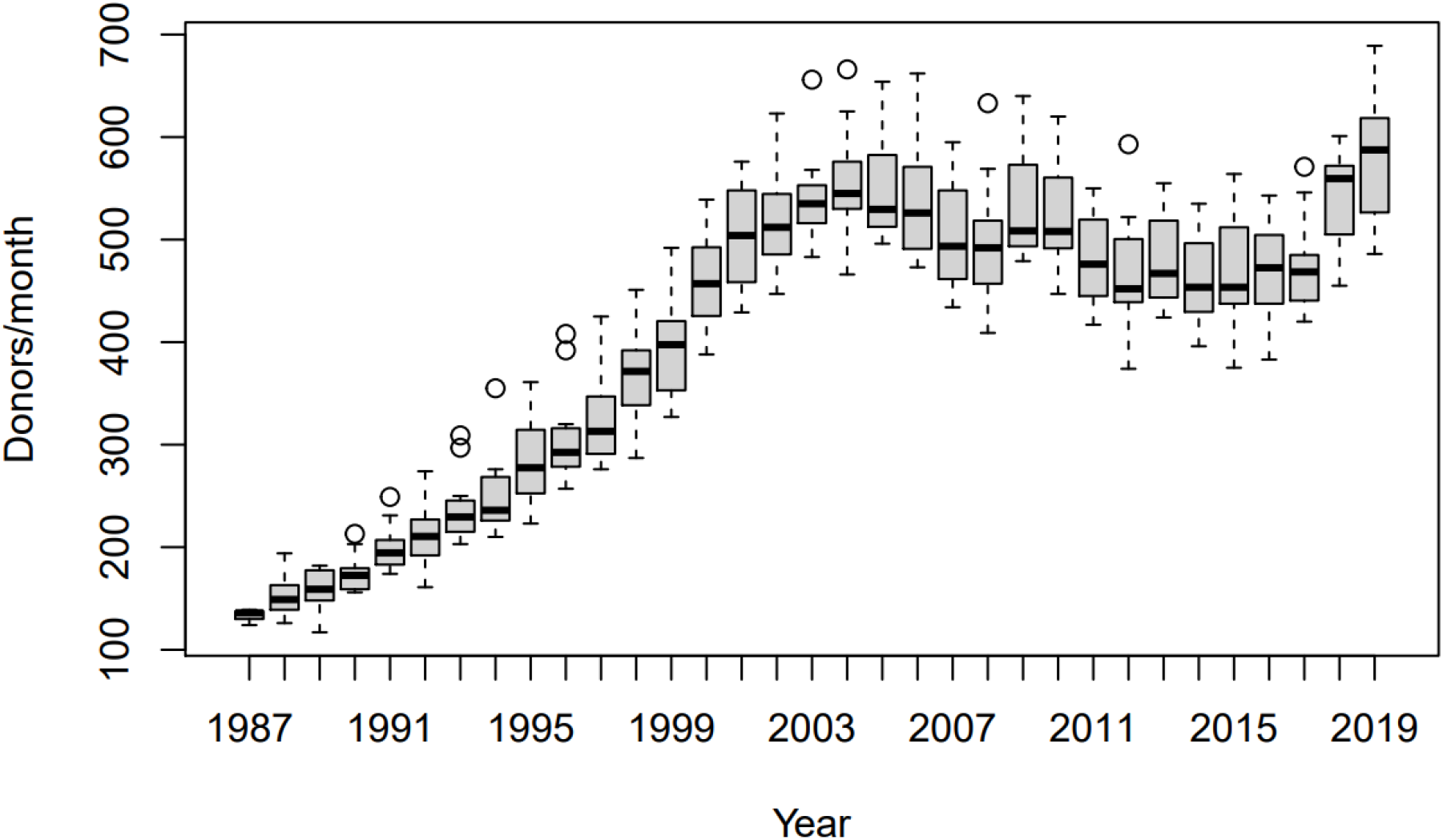
Monthly incidence of living kidney donation across time. Boxplots reflect the distribution of donations for the 12 months of each year. Bold line reflects the median, shaded box the interquartile range, and the tails 1.5 IQRs from the median, with outliers displayed as circles. Based on the median of each year, between-year donation increased exponentially from 1987-2004, declined from 2004-2016, and surged from 2017-2019. However, based on the interquartile range, month-to-month variation in donation substantially increased over time. The interclass-correlation coefficient comparing between-year vs. within-year variation decreased from 0.94 in the period 1987-2004 to 0.28 in the period 2005-2019.

These results were similar when stratifying for related and unrelated donation. For related donors, ICC was 0.93 prior to 2005 and 0.33 thereafter. Intra-year variation increased by 22% from 45 to 55. For unrelated donors, ICC was 0.92 prior to 2005 and 0.26 thereafter. Intra-year variation increased by 10% from 48 to 53 between these time periods.

### Within-Year Seasonal Variation in Living Kidney Donors

When looking at monthly trends in kidney donation, kidney donation was highest in the summer months—particularly June and July—for all subgroups. These trends have been historically stable, as donations have been higher in summer months (Figure 2). IRR was reported with January as the reference. Among related donors, IRRs for February were 1.05 [95% CI 0.94-1.18] for the highest income tertile, 1.00 [0.88-1.13] for the middle income tertile, and 1.00 [0.86-1.18] for the lowest income tertile. For March, IRRs were 1.01 [0.90-1.15], 0.98 [0.86-1.13], and 1.01 [0.85-1.19]. For April, IRRs were 1.00 [0.87-1.15], 0.95 [0.82-1.10], and 1.04 [0.89-1.22] relative to January. For May, IRRs were 1.04 [0.92-1.16], 1.00 [0.92-1.15], and 1.03 [0.89-1.20]. For June, IRRs were 1.34 [1.20-1.50], 1.25 [1.11-1.42], and 1.22 [1.05-1.43]. For July, IRRs were 1.27 [1.14-1.42], 1.20 [1.06-1.36], and 1.19 [1.01-1.39]. For August, IRRs were 1.15 [1.03-1.28], 1.05 [0.93-1.18], and 1.18 [1.01-1.39]. For September, IRRs were 0.96 [0.86-1.07], 0.98 [0.86-1.10], and 1.04 [0.87-1.23]. For October, IRRs were 1.05 [0.94-1.17], 1.01 [0.90-1.13], and 1.02 [0.87-1.20]. For November, IRRs were 1.09 [0.98-1.20], 1.04 [0.93-1.16], and 1.01 [0.86-1.20]. For December, IRRs were 1.08 [0.97-1.20], 1.03 [0.93-1.15], and 1.10 [0.95-1.28]. IRRs were statistically significant for all income tertiles in June, July, and August with the single exception of the middle income tertile in August (Figure 3A).

**Figure 2.**
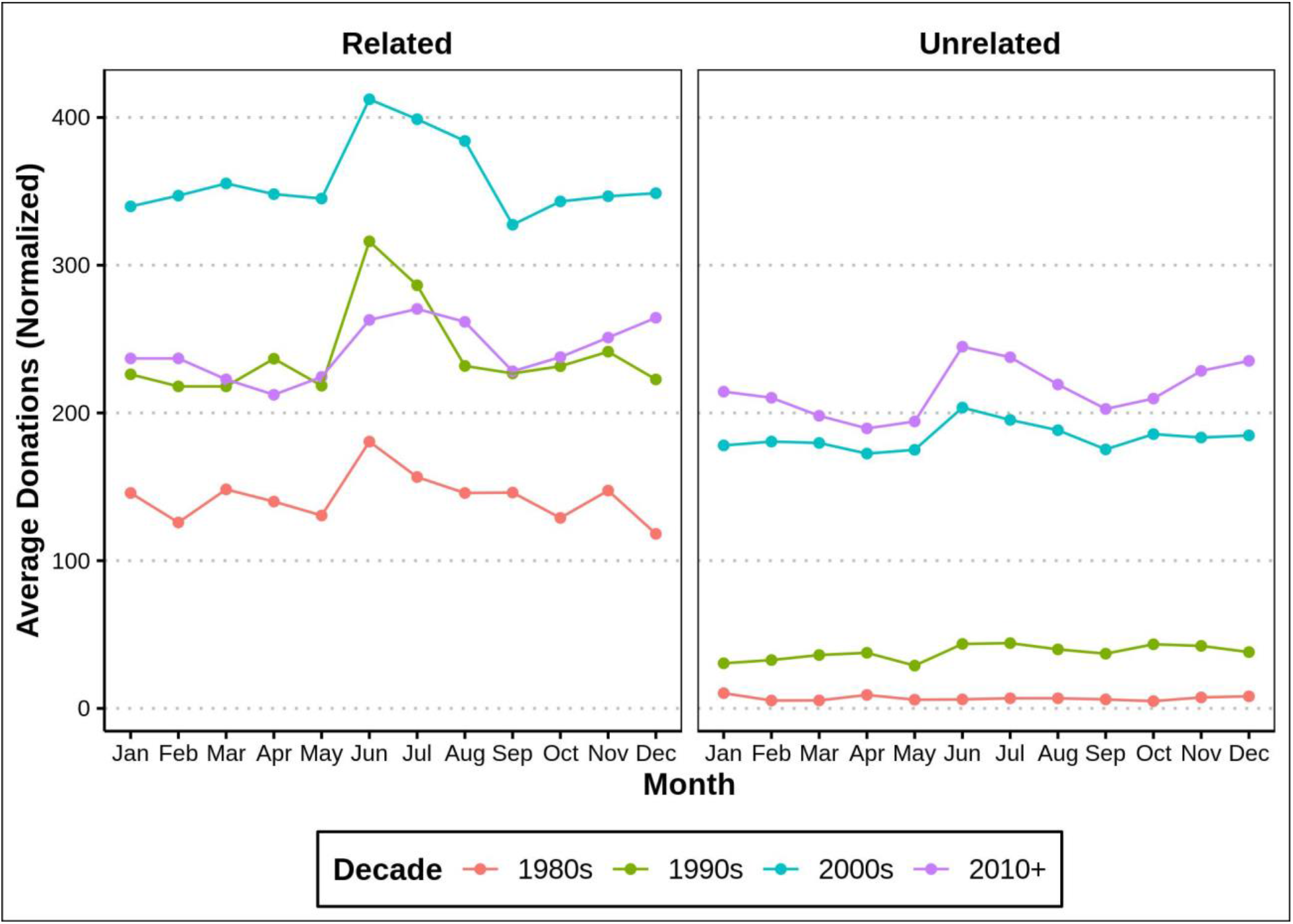
Average living kidney donations per month by decade for related and unrelated donors. Each data point reflects the mean number of donations in the month over the years of the labelled time period. Summer donations for related donors were 34 donations per month (25%) higher than non-summer donations in the 1980s, 52 (23%) higher in the 1990s, 54 (16%) higher in the 2000s, and 31 (13%) higher in the 2010s. For unrelated donors these numbers were 1 donation per month (14%) higher in the 1980s, 6 (17%) higher in the 1990s, 17 (10%) higher in the 2000s, and 35 (17%) higher in the 2010s.

**Figure 3A.**
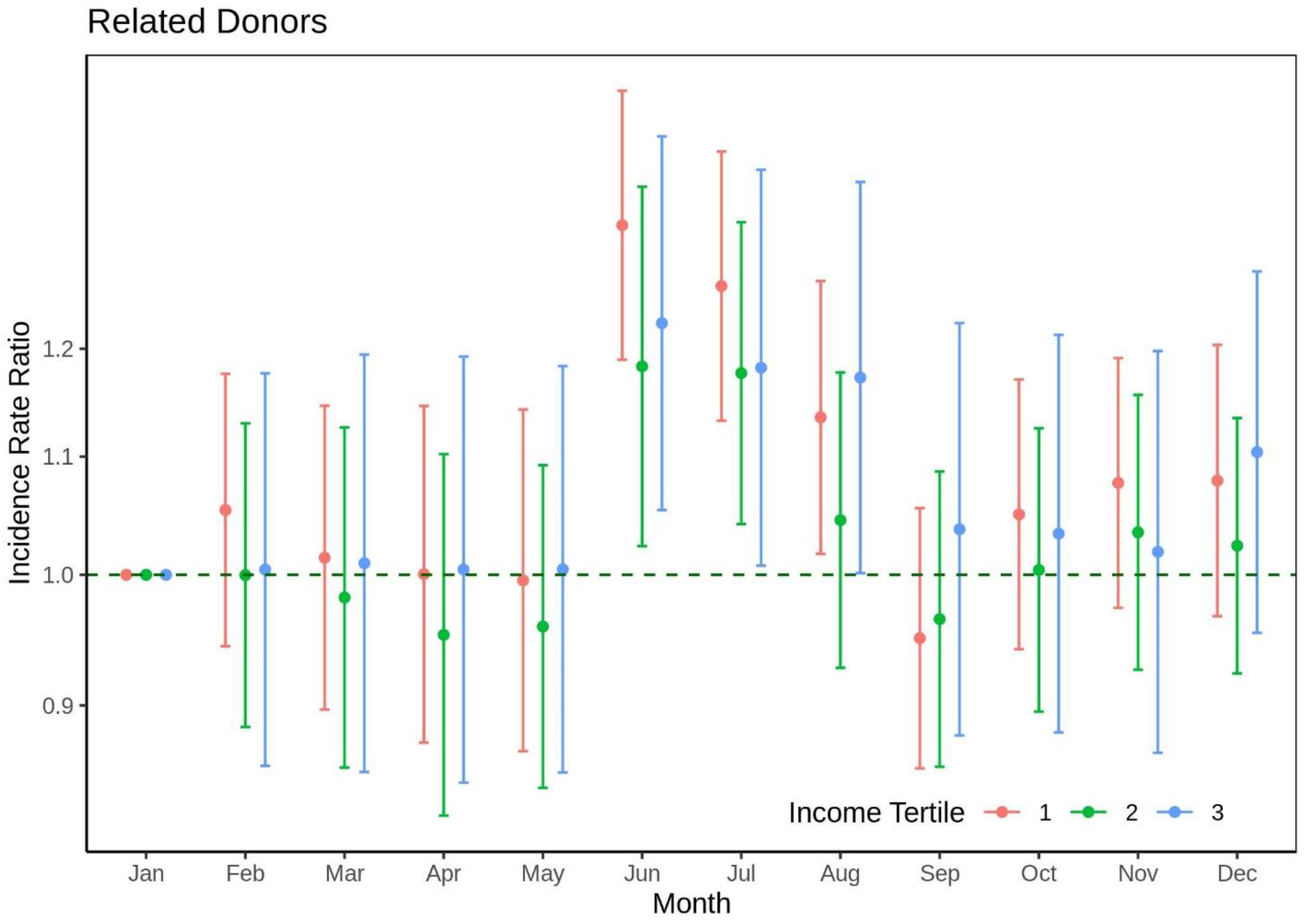
Incidence rate ratio of living kidney donation by biological relatives based on month of donation. The dot reflects the IRR relative to January and error bars represent the 95% confidence interval. Orange reflects the highest income tertile ($73,544+), green the middle tertile ($52,635-$73,544), and blue the lowest tertile (<$52,635).

Very similar patterns were observed for unrelated donors. Thus, IRRs for February were 1.00 [95% CI 0.81-1.24] for the highest income tertile, 0.94 [0.77-1.15] for the middle income tertile, and 1.03 [0.86-1.23] for the lowest income tertile. For March, IRRs were 0.98 [0.79-1.21], 0.97 [0.80-1.17], and 0.96 [0.81-1.14] for the income tertiles relative to January. For April, IRRs were 0.95 [0.76-1.19], 0.95 [0.78-1.15], and 0.99 [0.84-1.17] relative to January. For May, IRRs were 0.97 [0.77-1.21], 0.92 [0.75-1.12], and 1.02 [0.85-1.22]. For June, IRRs were 1.13 [0.92-1.40], 1.14 [0.93-1.40], and 1.19 [1.01-1.39]. For July, IRRs were 1.11 [0.90-1.38], 1.08 [0.88-1.33], and 1.17 [0.99-1.37]. For August, IRRs were 1.01 [0.82-1.26], 1.02 [0.84-1.25], and 1.12 [0.95-1.33]. For September, IRRs were 0.95 [0.77-1.17], 0.95 [0.77-1.16], and 1.03 [0.86-1.23]. For October, IRRs were 1.06 [0.86-1.31], 0.96 [0.80-1.15], and 1.07 [0.91-1.26]. For November, IRRs were 1.11 [0.90-1.37], 1.01 [0.83-1.24], and 1.07 [0.90-1.26]. For December, IRRs were 1.09 [0.89-1.35], 1.02 [0.82-1.26], and 1.09 [0.91-1.31]. The IRR was statistically significant for the lowest tertile in June (Figure 3B).

**Figure 3B.**
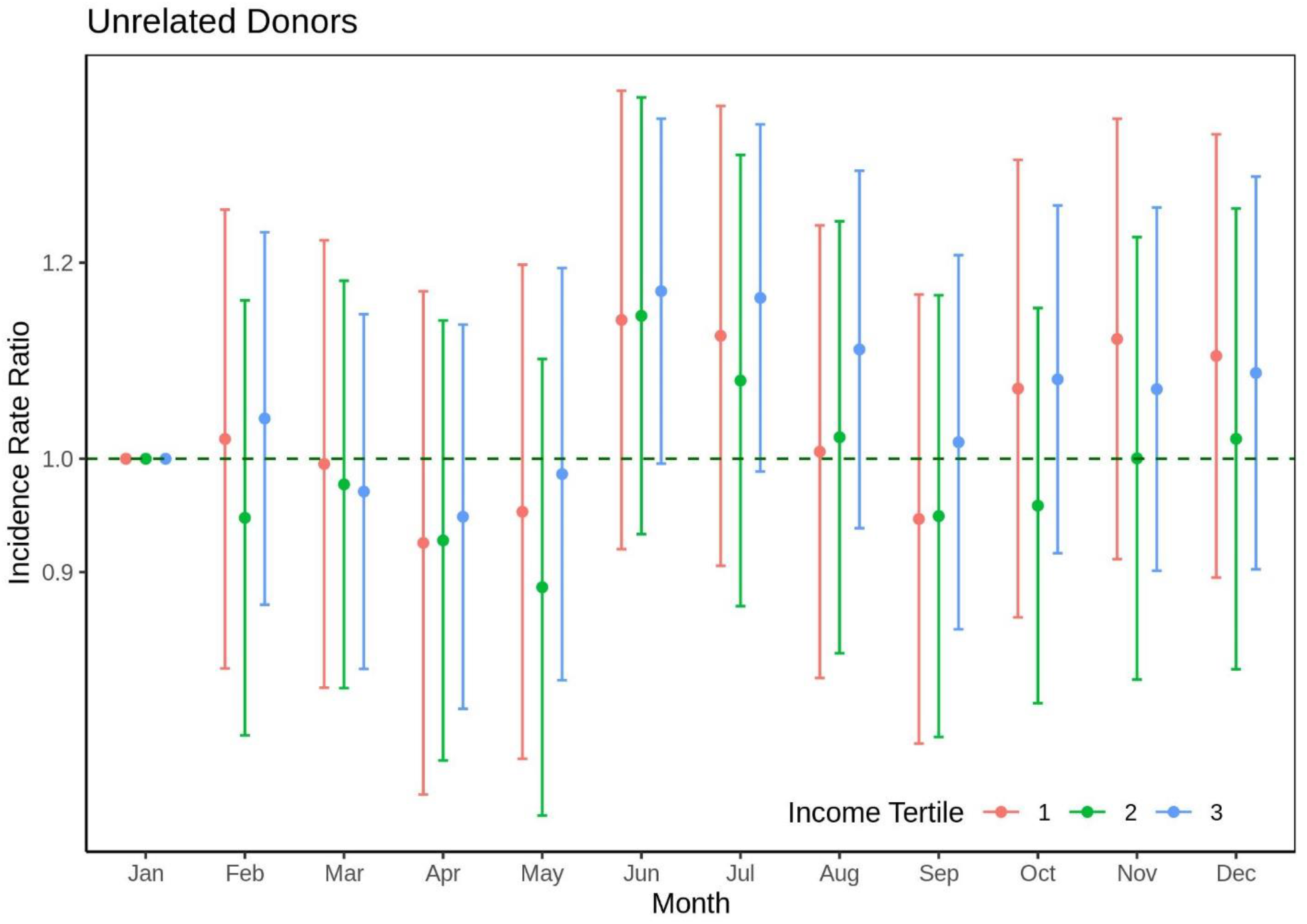
Incidence rate ratio of living kidney donation by nonrelatives based on month of donation. The dot reflects the IRR relative to January and error bars represent the 95% confidence interval. Orange reflects the highest income tertile ($73,544+), green the middle tertile ($52,635-$73,544), and blue the lowest tertile (<$52,635).

### Between-Year Seasonal Variation in Living Kidney Donors

Between-year trends demonstrate different trends between related and unrelated groups, with additional interaction with income group. For related donors, trends were uniformly positive for cases up to 2004 and uniformly negative for 2005-2019 (Supplemental Figure XX). For unrelated donors, slopes were uniformly positive for February and July in all income groups from 1996 to 2004 (Supplemental Figure XX). These more complex dynamics can be understood through linear regression which yields the rate of increase/decrease for a given month viewed across the years. Slopes varied by season and income group for unrelated donors from 2005 to 2013: In the highest income group, cases increased on average by 0.1 cases each February and increased 1.8 each July. In the middle-income group, cases decreased by 0.3 each February but increased by 2.3 each July. In the lowest income group, cases decreased by 0.7 cases each February and by 0.3 cases each July. After the second turning point, donations increased in all groups but again varied with income tertile and season. In the highest income group, cases increased on average by 3.4 cases each February and increased by 4.2 each July. In the middle-income group, cases increased by 1.6 each February and increased by 5.7 each July. In the lowest income group, cases increased by only 1.5 cases each February and by 0.8 cases each July. (Supplemental Figure XX, Supplemental Table XX).

## DISCUSSION

In this national study of trends in living kidney donation, the effects of seasonal variation over time were quantified. The ICC for seasonal donations changed from 0.94 before 2005 to 0.28 thereafter, while the within-year variation increased by 30%, from 40 to 52. These effects were more marked in related donors. For related donors, donation rates were 13-25 percent higher during summer months across all decades since the advent of living kidney donation; for unrelated donors donation rates were 10-17 percent higher. Using incidence rate ratios to quantify the increase while accounting for covariates, for related donors, June donations were 1.34-fold more frequent than January donation among those in the highest income tertile, 1.25-fold higher in the middle income tertile, and 1.22-fold higher in the lowest income tertile. For July these were 1.27-folder higher, 1.20-fold higher, and 1.19-fold higher. For unrelated donors, June donations were 1.13-fold higher, 1.14-fold higher, and 1.19-higher for the three income tertiles. For July, these were 1.11-fold higher, 1.08-fold higher, and 1.17-fold higher. Thus, non-summer months are associated with significantly lower rates of living kidney donation within a given year.

All prior studies of trends in living kidney donation have offered a narrative on annual changes in living donation. However, our study reveals that a narrative based only on year-to-year donation rates is most appropriate for the years 1987-2004 and less so for the years 2005-2019. During these latter years, particularly in unrelated donors, annual trends were mediated by seasonality of donation and donor income level. Furthermore, the within-year variance increased from the previous period and intra-class correlation plummeted. Taken together, these results indicate the rise of novel factors since 2005 that differentially impact potential donors depending on season of donation and socioeconomic status.

While these results reaffirm our previous report wherein we showed a decrease in related donors between 2005-2017 in addition to a slight increase in unrelated donors during that same window, our current study introduces seasonal variation as a phenomenon by itself and a modifier of previously reported historical trends in kidney donation. We posit that within-year seasonal variation in donation reflects barriers that all donor candidates face when they seek to donate to a relative or a non-relative, regardless of income level; such barriers subside in the summer while remaining present during the rest of the year. We further suggest that these barriers might relate to work-school calendars that allow much more freedoms to a potential donor candidate in the summer months than in the non-summer months. Our results indicate that structural barriers disproportionately affect lower-income households, a finding previously unreported in the literature.

This study has several limitations. First, using zip code data provides a crude picture of donor socioeconomic status because donors’ income may not align with their zip code’s median. Second, using aggregate count data eliminates many individual-level characteristics that may influence donation. We could not stratify the analysis by all plausible factors because of the limited statistical power inherent in such an approach. Granted, this study also has one key strength. We uncover at a national level that barriers to donation exist that may impact the less financially resilient donor candidate in non-summer months of any given year, and even more so in non-summer months during a national-wide financial crisis. That the barriers are most evident when annual trends are viewed from the perspective of non-summer months points to indirect costs linked to the impact of donation on school-work schedules. These inferences suggest actionable way forward for future programs to promote donation: further studies will be needed to identify the details of these structural barriers and develop specific policy recommendations to bypass the disincentives to donation.

In conclusion, we have suggested seasonality as a modifier for future studies of trends in living kidney donation, as we observed that non-summer donation rates are much lower than summer rates. That these seasonal trends persist across income groups even during times of economic downturn indicates that broad structural forces contribute to trends in seasonality. Overcoming the indirect costs and other barriers associated with non-summer donations has the potential to increase non-summer donations.

## Data Availability

All data produced in the present work are contained in the manuscript

## Abbreviations

(LKD: Living kidney donor
(ESRD): End-stage renal disease

## ACKNOWLEDGEMENTS

This work was supported by grant number K24AI144954 (Segev) from the National Institute of Allergy and Infectious Diseases (NIAID) and K08AG065520-01 (Muzaale) from the National Institute on Aging. The analyses described here are the responsibility of the authors alone and do not necessarily reflect the views or policies of the Department of Health and Human Services, nor does mention of trade names, commercial products or organizations imply endorsement by the U.S. Government.

The data reported here have been supplied by the Hennepin Healthcare Research Institute (HHRI) as the contractor for the Scientific Registry of Transplant Recipients (SRTR). The interpretation and reporting of these data are the responsibility of the author(s) and in no way should be seen as an official policy of or interpretation by the SRTR or the U.S. Government.

